# The increase in paraoxonase-1 is associated with a decrease in left ventricular volume in kidney transplant recipients

**DOI:** 10.1101/2020.01.28.20018655

**Authors:** Philip W. Connelly, Andrew T. Yan, Michelle M. Nash, Rachel Wald, Charmaine Lok, Lakshman Gunaratnam, Anish Kirpalani, G.V. Ramesh Prasad

**Author notes:** Corresponding author: Philip W. Connelly, Keenan Research Centre for Biomedical Science of St. Michael’s Hospital, Room 413, 209 Victoria Street, Toronto, ON, Canada, M5B 1T8.

## Abstract

**Background:** Patients on dialysis have impaired cardiac function, in part due to increased fluid volume and ventricular stress. Restored kidney function through transplantation reduces left ventricular volume in both systole and diastole. Paraoxonase 1 (PON1) is reduced in patients on dialysis, which may be related to their impaired cardiac function. We tested the hypothesis that change in PON1 is associated with changes in left ventricular (LV) end-volume and LV mass after kidney transplantation.

**Methods:** Patients were studied before and 12 months after kidney transplantation. The control group was patients on dialysis not expected to receive a transplant in the next 12 months. Cardiac magnetic resonance imaging was used to measure LV end-diastolic and end-systolic volume and LV mass. PON1 was measured by arylesterase activity and by mass.

**Results:** PON1 mass and activity were not different between the groups at baseline. Both PON1 mass and activity were increased post-kidney transplantation (p<0.0001 for change). The change in PON1 mass (p=0.0062) and PON1 arylesterase activity (p=0.0254) were inversely correlated with the change in NT-proBNP for patients receiving a kidney transplant. However, only the change in the PON1 mass, but not the change in PON1 arylesterase, was inversely correlated with the change in left ventricular volume (ml/m^2.7^) (p=0.0146 and 0.0114 for diastolic and systolic, respectively) and with the change in hemoglobin (p=0.0042).

**Conclusions:** PON1 mass and activity increase after kidney transplantation. The increase in PON1 mass is consistent with a novel relationship to the increase in hemoglobin and the decrease in LV end-systolic and end-diastolic volume.

## Introduction

Paraoxonase 1 (PON1) is a 43kDa protein synthesized primarily by the liver and released by hepatocytes to high density lipoproteins (HDL), to which it binds by an uncleaved N-terminal leader sequence. PON1 has both metal binding and hydrolase properties, functioning as an arylesterase and a hydrolase for paraoxon and chlorpyriphos oxon, respectively Sequence homology revealed a native function for PON1 as a lactonase [1, 2].

Kidney function is an important determinant of lipoprotein concentrations [3]. However, the amount of PON1 activity seen in states of reduced kidney function, including end-stage kidney disease (ESKD) requiring hemodialysis (HD) or peritoneal dialysis (PD), and partially restored kidney function, such as after kidney transplantation, remains to be fully established.

The first report that PON1 activity was reduced in patients on hemodialysis was by Schiavon et al. [4]. Hasselwander et al. [5], who also found both paraoxonase and arylesterase were lower in patients on dialysis. We observed lower PON1 in patients on standard hemodialysis and home nocturnal hemodialysis that was inversely correlated with C-reactive protein (CRP), a marker of inflammation [6]. In a cross-sectional study comparing a control group with patients on HD and patients with a kidney transplant, Sztanek et al. [7] found PON1 activity (measured as paraoxonase or lactonase) to be lowest for HD patients, followed by those with kidney transplant and highest in control subjects. Samouilidou et al. [8] measured PON1 mass and found it to be markedly reduced in HD patients compared to controls and patients with CKD.

Bhattacharyya et al. [9] reported that PON1 Q192R genotype, paraoxonase, and arylesterase activities were related to cardiovascular outcomes in a cohort of subjects undergoing elective diagnostic cardiac catheterization. The lowest quartile of arylesterase activity had a 4.5-fold hazard ratio for nonfatal myocardial infarction or cerebrovascular accident. Hammadah et al. [10] reported event-free survival over five years for subjects with chronic heart failure from the Atlanta Cardiomyopathy Consortium. BNP was highest, and HDL cholesterol was lowest for the lowest tertile of arylesterase activity. The lowest tertile of baseline arylesterase activity was associated with a 2.6 hazard ratio for heart failure events (death, cardiac transplant, ventricular assist device).

Patients with chronic kidney disease (CKD) are at increased risk for cardiovascular disease, particularly nonatherosclerotic disease characterized by left ventricular hypertrophy [11]. Cardiac magnetic resonance imaging (CMR) is particularly useful as a noninvasive approach to measure left ventricular volume and mass due to its accuracy and precision [12]. We have recently reported the left ventricular changes in patients on dialysis receiving a kidney transplant. While both left ventricular volume and adiponectin levels decreased with a kidney transplant, the change in adiponectin level and the change in HDL cholesterol were not associated with the change in left ventricular volume [13]. CMR is, therefore, a useful tool to correlate biomarker changes after transplantation with measurable changes in cardiac function.

In summary, the majority of studies of patients on dialysis have measured PON1 enzymatic activity. There is inconsistency in the reports, but the consensus is that activity is lowest for patients on dialysis, higher for patients that have received a kidney transplant, and highest for normal control subjects. This gradient in PON1 enzymatic activity is consistent with the gradient of kidney function. However, these studies were all cross-sectional in design. No study to date has prospectively determined the change in PON1 mass or activity with kidney transplantation, a procedure that partially restores kidney function so that patients with ESKD no longer require dialysis. Further, there has been no study that compared the change in PON1 and the change in left ventricular indices.

## Materials and Methods

The recruitment of participants for this study has been described [14, 13]. The study sample size target of 42 subjects per group was designed to detect a 5 µg/ml change in adiponectin, with expected attrition of 20%. The methods for CMR measurement of left ventricular volume and left ventricular mass have been described in detail [14]. Briefly, CMR was performed with a 1.5-tesla whole-body magnetic resonance scanner (Intera: Philips Medical Systems, Best, The Netherlands) using a phased-array cardiac coil and retrospective vectorographic gating. All CMR studies were analyzed by readers blinded to the information about the patient and the time point of data acquisition. CMR data were analyzed by an experienced reader using cvi42 software (Circle Cardiovascular, Calgary, Canada). Left ventricular volumes and mass were allometrically adjusted by dividing by height in meters^2.7^ [15]. Adiponectin was measured using the Meso Scale Discovery human adiponectin assay (#K151BXC-2, Meso Scale Diagnostics, Rockville, Maryland) and calibrated to the Millipore enzyme-linked immunoassay (#EZHADP-61-K, Millipore, St. Charles, MO, USA). N-terminal proB-type natriuretic peptide was measured on the Roche Cobas 6000 601e module (Mississauga, ON, Canada). Measurement of PON1 was an a priori secondary variable. PON1 activity as hydrolysis of phenyl acetate and as enzyme mass were measured as described [16, 17]. PON1 phenotype was determined by the two substrate method of Richter et al. [18].

Statistical analysis was done using SAS version 9.4 (Cary, NC, USA). Graphs were plotted using GraphPad Prism vs 8.0). Between-group comparisons were made using unpaired t-test.

The study was approved by the Research Ethics Board at St. Michael’s Hospital (REB 10-239) and by the ethics boards at the collaborating sites. All subjects provided written informed consent. The work described has been carried out in accordance with The Code of Ethics of the World Medical Association (Declaration of Helsinki) for experiments involving humans.

## Results

The characteristics of the study population are shown in Table 1. Hemoglobin and albumin concentrations at baseline were significantly different between the participants on peritoneal dialysis versus hemodialysis. Therefore, a sensitivity analysis was performed, and the univariate analyses are presented separately by baseline dialysis type.

**Table 1.** Characteristics of study subjects by treatment group and baseline dialysis modality.

|  | Hemodialysis at baseline |  |  | Peritoneal dialysis at baseline |  |  |
| --- | --- | --- | --- | --- | --- | --- |
|  | Dialysis | Transplant |  | Dialysis | Transplant |  |
| Women/Men | 8/23 | 10/17 |  | 4/8 | 2/10 |  |
| Age, yrs | 54.1 ± 9.6 | 46.3 ± 12.1 | 0.008 | 59.2 ± 13.8 | 47.1 ± 13.7 | 0.04 |
| **Baseline** |  |  |  |  |  |  |
| Serum Creatinine $\mu\text{mol/L}$ | 674 ± 229 | 694 ± 230 | 0.75 | 789 ± 199 | 926 ± 332 | 0.23 |
| BMI, $\text{kg/m}^2$ | 27.2 ± 5.2 | 25.5 ± 4.9 | 0.2 | 25.7 ± 4.1 | 27.1 ± 3.6 | 0.39 |
| Hemoglobin, g/L | 118.7 ± 15.3<sup>^</sup> | 121.5 ± 13.4<sup>*</sup> | 0.46 | 109.3 ± 11.7 | 106.7 ± 17.0 | 0.67 |
| Total Chol, mM | 4.0 ± 1.3 | 4.16 ± 1.22 | 0.63 | 4.15 ± 1.04 | 4.21 ± 1.11 | 0.89 |
| Triglycerides, mM | 1.46 ± 0.85 | 1.93 ± 1.39 | 0.12 | 1.53 ± 0.53 | 2.03 ± 0.84 | 0.10 |
| LDL Chol, mM | 2.17 ± 1.12 | 2.17 ± 0.97 | 0.98 | 2.35 ± 1.0 | 2.38 ± 0.95 | 0.93 |
| HDL Chol, mM | 1.19 ± 0.33 | 1.13 ± 0.32 | 0.51 | 1.11 ± 0.36 | 0.90 ± 0.19 | 0.10 |
| Albumin, g/L | 41.6 ± 4.2<sup>#</sup> | 41.7 ± 3.8<sup>§</sup> | 0.91 | 35.8 ± 2.4 | 36.9 ± 3.5 | 0.40 |
| CRP, mg/L | 6.5 ± 10.2 | 3.8 ± 5.0 | 0.21 | 6.5 ± 7.3 | 6.0 ± 6.0 | 0.88 |
| LV systolic volume, $\text{ml/m}^{2.7}$ | 15.9 ± 5.5 | 17.5 ± 6.5 | 0.33 | 15.2 ± 5.0 | 19.3 ± 5.8 | 0.08 |
| LV diastolic volume, $\text{ml/m}^{2.7}$ | 39.2 ± 10.2 | 40.6 ± 10.7 | 0.63 | 37.0 ± 7.8 | 45.0 ± 9.4 | 0.036 |
| LV mass index, $\text{g/m}^{2.7}$ | 30.8 ± 9.9 | 29.2 ± 8.6 | 0.50 | 28.7 ± 4.5 | 31.4 ± 8.2 | 0.32 |
| Ln NT-proBNP, pg/ml | 7.49 ± 1.55 | 7.23 ± 1.27 | 0.50 | 6.81 ± 1.38 | 7.30 ± 1.37 | 0.41 |
| Adiponectin, $\mu\text{g/ml}$ | 23.0 ± 13.5 | 20.6 ± 11.0 | 0.47 | 20.4 ± 10.7 | 27.5 ± 10.8 | 0.13 |
| PON1 $\mu\text{g/ml}$ | 98.2 ± 24.2 | 92.0 ± 22.9 | 0.32 | 80.8 ± 19 | 79.5 ± 26 | 0.89 |
| PON1 arylesterase, U/ml | 88.1 ± 19.2 | 82.3 ± 19.5 | 0.25 | 82.2 ± 19.5 | 78.4 ± 19.7 | 0.65 |
| PON1 specific activity, U/$\mu\text{g}$ | 0.91 ± 0.16 | 0.91 ± 0.18 | 0.99 | 1.02 ± 0.11 | 1.02 ± 0.18 | 0.92 |

| **Month 12** |  |  |  |  |  |  |
| --- | --- | --- | --- | --- | --- | --- |
| Serum creatinine $\mu\text{mol/L}$ | 757 ± 234 | 129 ± 103 | <0.0001 | 696 ± 177 | 118 ± 31 | <0.0001 |
| BMI, $\text{kg/m}^2$ | 27.6 ± 5.7 | 26.7 ± 4.7 | 0.49 | 25.0 ± 3.4 | 28.2 ± 5.4 | 0.11 |
| Hemoglobin, g/L | 116.6 ± 12.1 | 134.3 ± 20.3 | 0.0003 | 113.1 ± 13.3 | 137.6 ± 18.6 | 0.0012 |
| Total Chol, mM | 3.98 ± 1.32 | 4.39 ± 1.13 | 0.21 | 3.94 ± 1.07 | 4.33 ± 1.17 | 0.40 |
| Triglycerides, mM | 1.56 ± 0.96 | 1.85 ± 1.38 | 0.34 | 1.54 ± 0.58 | 1.90 ± 0.87 | 0.24 |
| LDL Chol, mM | 2.08 ± 0.93 | 2.33 ± 0.81 | 0.29 | 2.18 ± 0.90 | 2.14 ± 0.72 | 0.90 |
| HDL Chol, mM | 1.13 ± 0.37 | 1.24 ± 0.39 | 0.29 | 1.05 ± 0.29 | 1.19 ± 0.28 | 0.25 |
| Albumin, g/L | 42.8 ± 3.5 | 43.6 ± 2.9 | 0.36 | 36.5 ± 2.75 | 43.5 ± 3.0 | <0.0001 |
| CRP, mg/L | 4.1 ± 3.9 | 3.0 ± 4.0 | 0.32 | 9.7 ± 13.3 | 4.8 ± 4.9 | 0.26 |
| LV systolic volume, $\text{ml/m}^{2.7}$ | 15.5 ± 5.2 | 14.5 ± 4.4 | 0.47 | 15.4 ± 11.4 | 14.7 ± 12.8 | 0.72 |
| LV diastolic volume, $\text{ml/m}^{2.7}$ | 39.6 ± 10.4 | 36.9 ± 8.1 | 0.29 | 37.3 ± 29.5 | 37.1 ± 34.6 | 0.96 |
| LV mass index, $\text{g/m}^{2.7}$ | 30.2 ± 9.5 | 28.0 ± 6.0 | 0.29 | 28.5 ± 8.9 | 27.4 ± 4.5 | 0.70 |
| Ln NT-proBNP, pg/ml | 7.74 ± 1.53 | 4.81 ± 1.28 | <0.0001 | 7.40 ± 1.77 | 4.95 ± 1.25 | 0.0007 |
| Adiponectin $\mu\text{g/ml}$ | 21.7 ± 13.4 | 15.1 ± 7.1 | 0.021 | 23.3 ± 17.2 | 18.1 ± 11.3 | 0.39 |
| PON1 $\mu\text{g/ml}$ | 96.2 ± 21.1 | 106.8 ± 22.4 | 0.07 | 83.0 ± 17.1 | 100.3 ± 23.8 | 0.06 |
| PON1 arylesterase, U/ml | 88.1 ± 19.9 | 99.2 ± 19.5 | 0.038 | 82.5 ± 17.0 | 95.4 ± 26.4 | 0.18 |
| PON1 specific activity, U/$\mu\text{g}$ | 0.93 ± 0.2 | 0.95 ± 0.16 | 0.80 | 1.00 ± 0.13 | 0.95 ± 0.19 | 0.46 |

Baseline hemoglobin: \* $p=0.006$ for hemodialysis vs peritoneal dialysis for the transplant group; ^ $p=0.06$ for hemodialysis vs peritoneal dialysis for the dialysis group; Baseline albumin: # $p=0.0011$ for hemodialysis vs peritoneal dialysis for the transplant group; § $p<0.0001$ for hemodialysis vs peritoneal dialysis for the dialysis group.

Within dialysis type, baseline values for the dialysis versus transplant subjects were not significantly different, with the exception of a lower left ventricular volume at diastole (ml/m^2.7^) for the peritoneal dialysis subjects. Allometrically adjusted left ventricular mass (g/m^2.7^) was not different between the two groups.

Kidney transplantation resulted in similar, statistically significant increases in hemoglobin concentration for both dialysis groups. Of note, kidney transplantation did not significantly affect albumin in the hemodialysis group, whereas there was a significant increase in albumin in the kidney transplant group that had been on peritoneal dialysis. The C-reactive protein concentration was not significantly different between the groups and was not significantly affected by kidney transplantation.

Univariate analyses of PON1 mass and activity comparing the dialysis and transplant groups are shown in Table 2. Baseline values were not significantly different between the two groups. Month 12 values for PON1 activity (p=0.0125) and PON1 mass (p=0.0119) were significantly higher in the transplant group, especially when analyzed as the change in activity or mass (p<0.0001). Notably, the standard deviation for the dialysis group and the transplant group was similar.

**Table 2.**
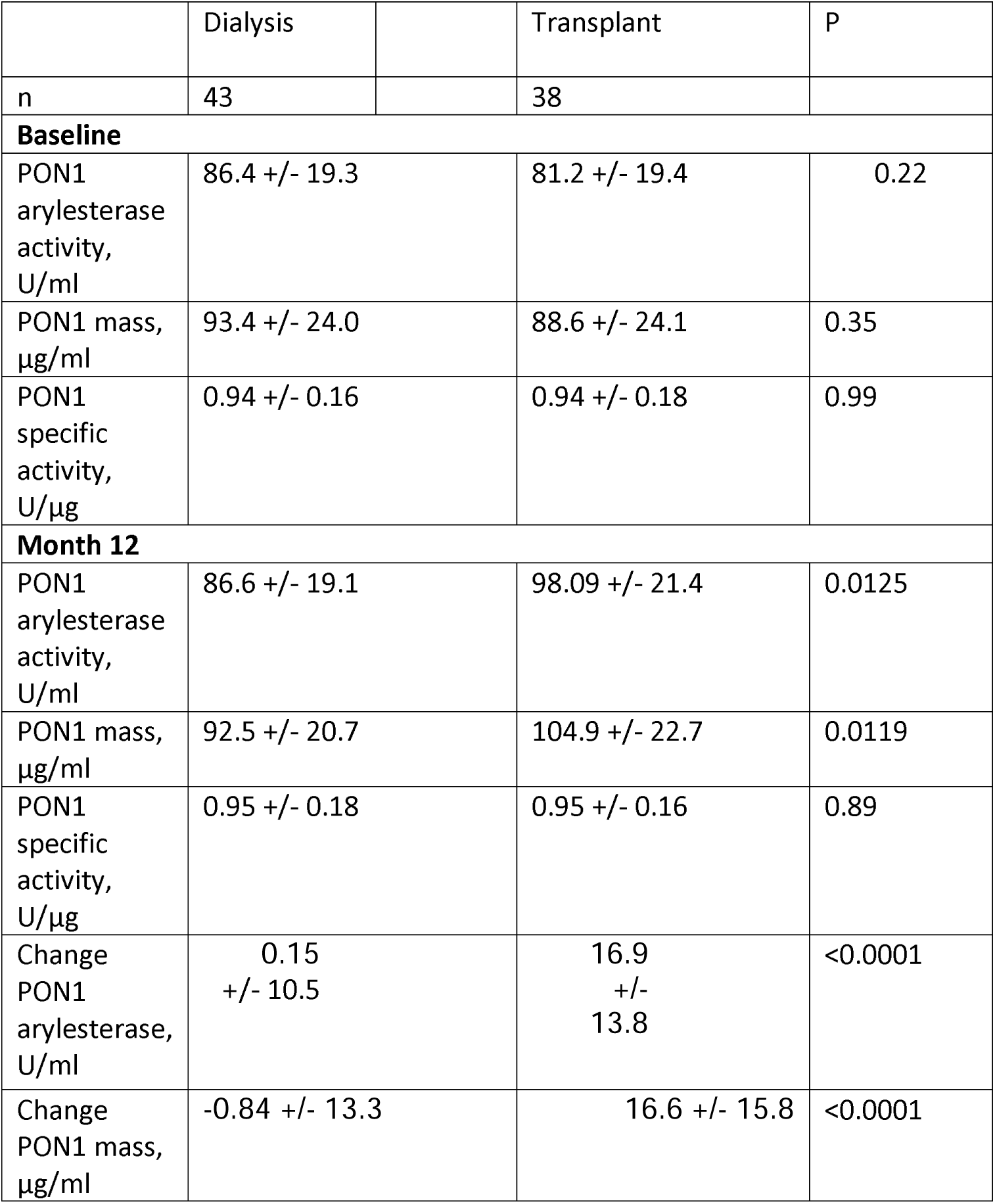
Paraoxonase 1 mass and activity by treatment group.

|  | Dialysis |  | Transplant | P |
| --- | --- | --- | --- | --- |
| n | 43 |  | 38 |  |
| **Baseline** |  |  |  |  |
| PON1<br>arylesterase<br>activity,<br>U/ml | 86.4 +/- 19.3 |  | 81.2 +/- 19.4 | 0.22 |
| PON1 mass,<br>µg/ml | 93.4 +/- 24.0 |  | 88.6 +/- 24.1 | 0.35 |
| PON1<br>specific<br>activity,<br>U/µg | 0.94 +/- 0.16 |  | 0.94 +/- 0.18 | 0.99 |
| **Month 12** |  |  |  |  |
| PON1<br>arylesterase<br>activity,<br>U/ml | 86.6 +/- 19.1 |  | 98.09 +/- 21.4 | 0.0125 |
| PON1 mass,<br>µg/ml | 92.5 +/- 20.7 |  | 104.9 +/- 22.7 | 0.0119 |
| PON1<br>specific<br>activity,<br>U/µg | 0.95 +/- 0.18 |  | 0.95 +/- 0.16 | 0.89 |
| Change<br>PON1<br>arylesterase,<br>U/ml | 0.15<br>+/- 10.5 |  | 16.9<br>+/-<br>13.8 | <0.0001 |
| Change<br>PON1 mass,<br>µg/ml | -0.84 +/- 13.3 |  | 16.6 +/- 15.8 | <0.0001 |

The most striking impact of dialysis modality was on the change in albumin, which was highly significant for subjects on peritoneal dialysis (p=0.0005), but not significant for subjects on hemodialysis (p=0.34) (Table 3). The change in systolic and diastolic left ventricular volume was greater for subjects on peritoneal dialysis and reached statistical significance in spite of the smaller sample size. The decrease in both measures for subjects on hemodialysis was consistent with a beneficial, albeit smaller, effect of kidney transplantation for this group. The change in PON1 activity and mass was significant for both dialysis modalities. The change in hemoglobin was also significant for both groups, although it was greater for the peritoneal dialysis subjects. The change in HDL cholesterol was also found to be significant for subjects in both dialysis groups. Thus, among parameters that are products of the liver, albumin was distinguished from PON1 and HDL cholesterol as having a strong effect due to dialysis type.

**Table 3.** Change in paraoxonase-1 mass and activity and relationship with hemoglobin and albumin by baseline dialysis modality

|  | Hemodialysis at baseline |  |  | Peritoneal dialysis at baseline |  |  |
| --- | --- | --- | --- | --- | --- | --- |
|  | Dialysis | Transplant |  | Dialysis | Transplant |  |
| n | N=28 | N=26 |  | 12 | 11 |  |
| Change in albumin, g/L | 1.04 ± 3.81 | 2.11 ± 4.41 | 0.34 | 0.67 ± 2.74 | 6.9 ± 4.4 | 0.0005 |
|  | N=30 | N=27 |  | N=12 | N=11 |  |
| Change in LV systolic volume, ml/m<sup>2.7</sup> | -0.43 ± 4.09 | -2.93 ± 5.91 | 0.067 | 0.28 ± 5.1 | -4.42 ± 4.76 | 0.033 |
| Change in LV diastolic volume, ml/m<sup>2.7</sup> | 0.28 ± 9.3 | -3.63 ± 7.98 | 0.096 | 0.33 ± 9.2 | -7.91 ± 9.45 | 0.046 |
|  | N=31 | N=27 |  | N=12 | N=11 |  |
| Change in PON1 activity, U/ml | 0.07 ± 11.0 | 16.9 ± 13 | <0.0001 | 0.35 ± 9.46 | 16.9 ± 16.2 | 0.0065 |
|  | N=31 | N=27 |  | N=12 | N=11 |  |
| Change in PON1 mass, µg/ml | -2.01 ± 12.9 | 14.9 ± 16.9 | <0.0001 | 2.2 ± 14.6 | 20.8 ± 12.32 | 0.0036 |
| Change PON1 specific activity, U/µg | 0.019 ± 0.12 | 0.03 ± 0.09 | 0.69 | -0.018 ± 0.1 | -0.061 ± 0.14 | 0.4 |
|  | N=31 | N=27 |  | N=12 | N=12 |  |
| Change in Hemoglobin, g/L | -2.1 ± 16.8 | 12.8 ± 23.2 | 0.0066 | 3.8 ± 12.1 | 30.9 ± 29.7 | 0.0106 |
| Change HDL cholesterol, mM | -0.05 ± 0.22 | 0.16 +/- 0.29 | 0.0081 | -0.05 ± 0.18 | 0.29 ± 0.3 | 0.0026 |

Correlation analysis is presented separately for the transplant group (Table 4) and the dialysis group (Table 5).

**Table 4.**
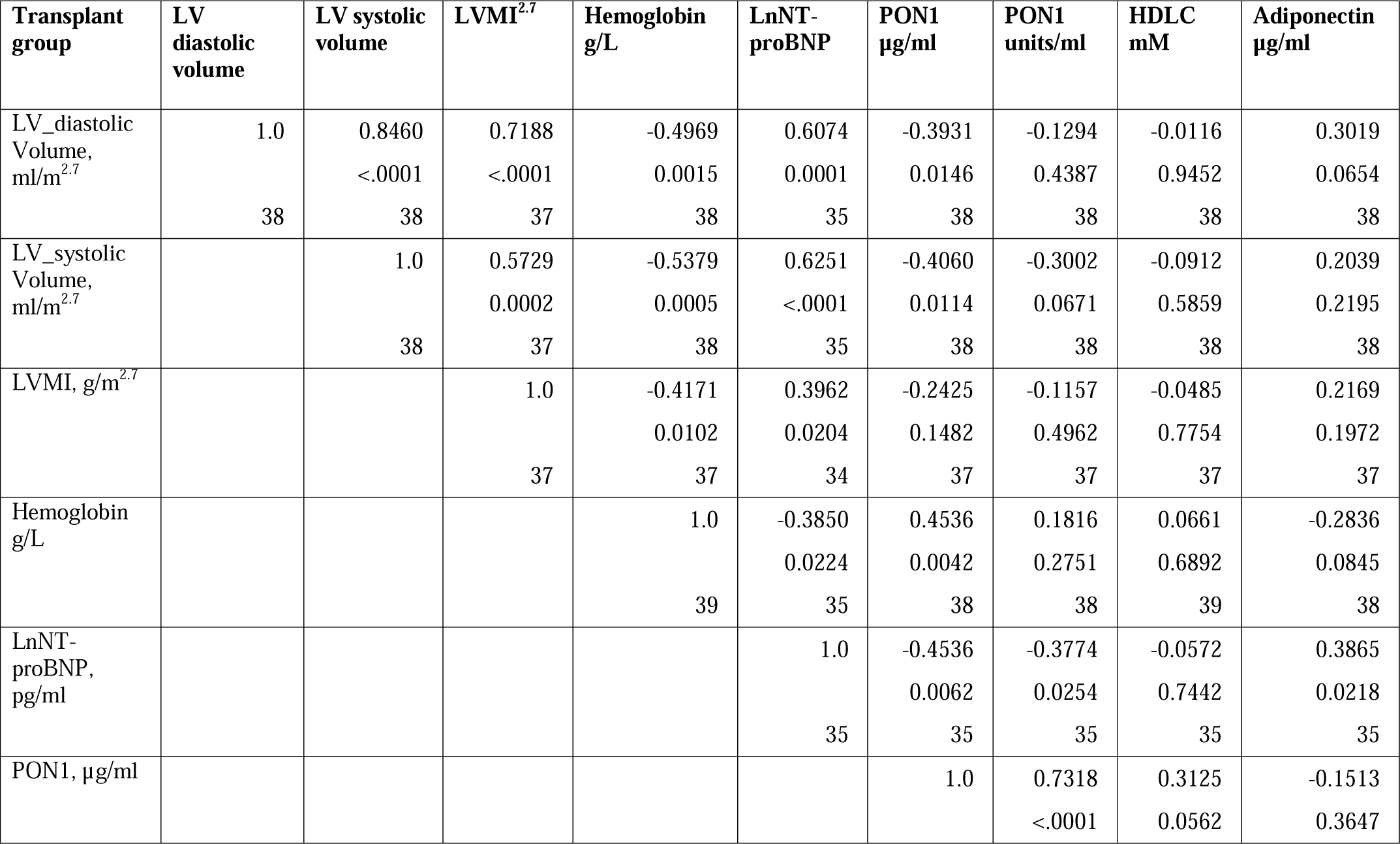
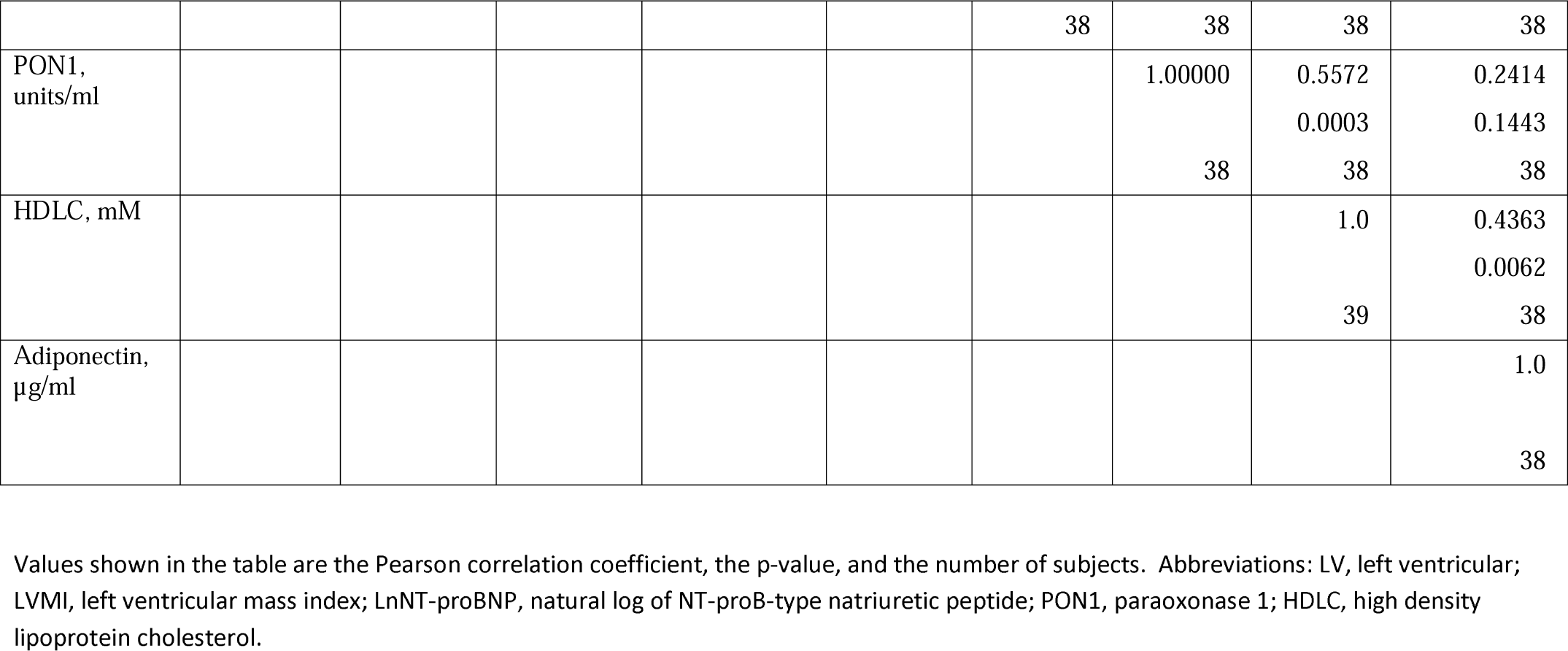
Pearson correlation of change in left ventricular measurements and hemoglobin with PON1, HDL cholesterol, and adiponectin for subjects receiving a kidney transplant.

**Table 5.**
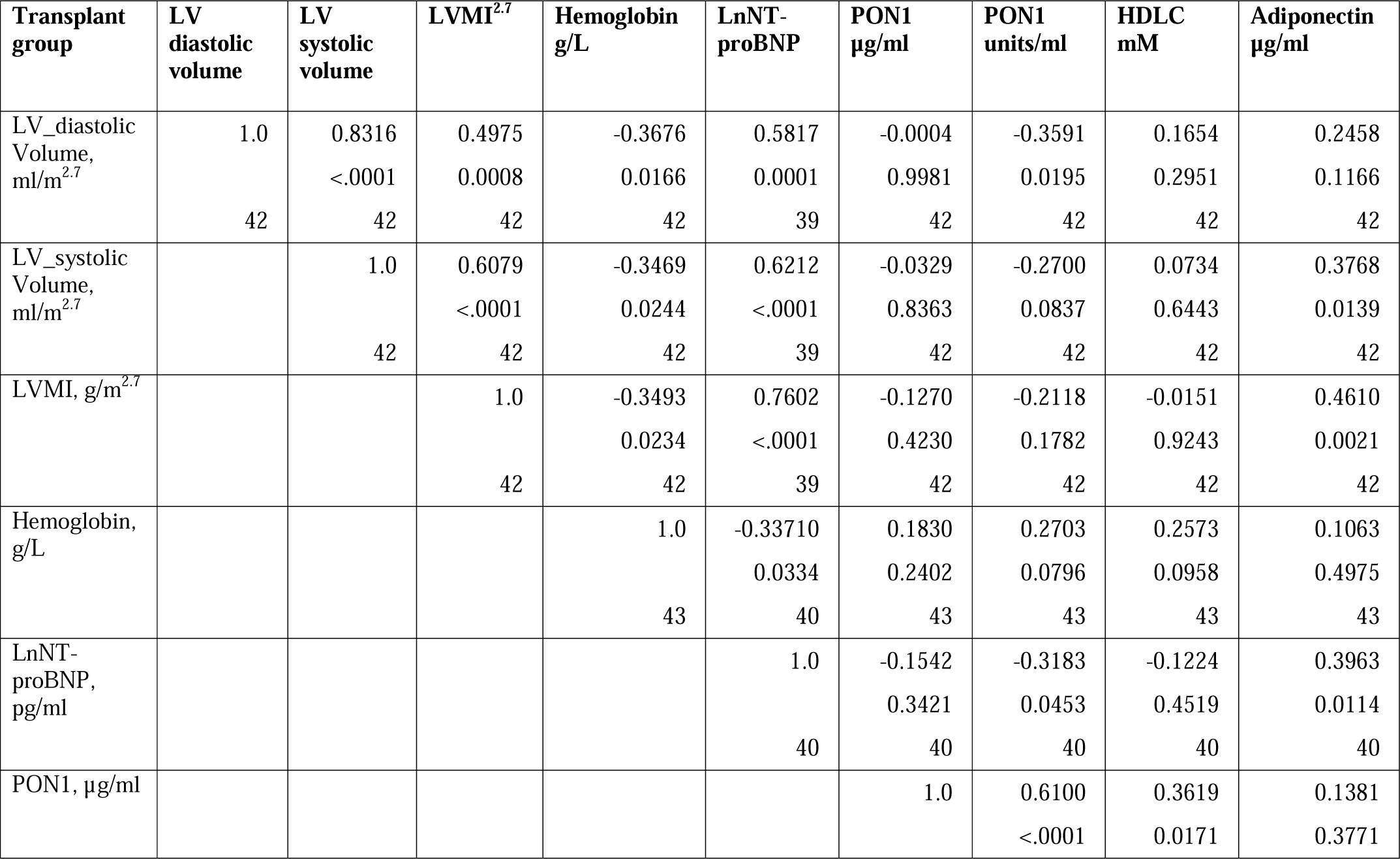
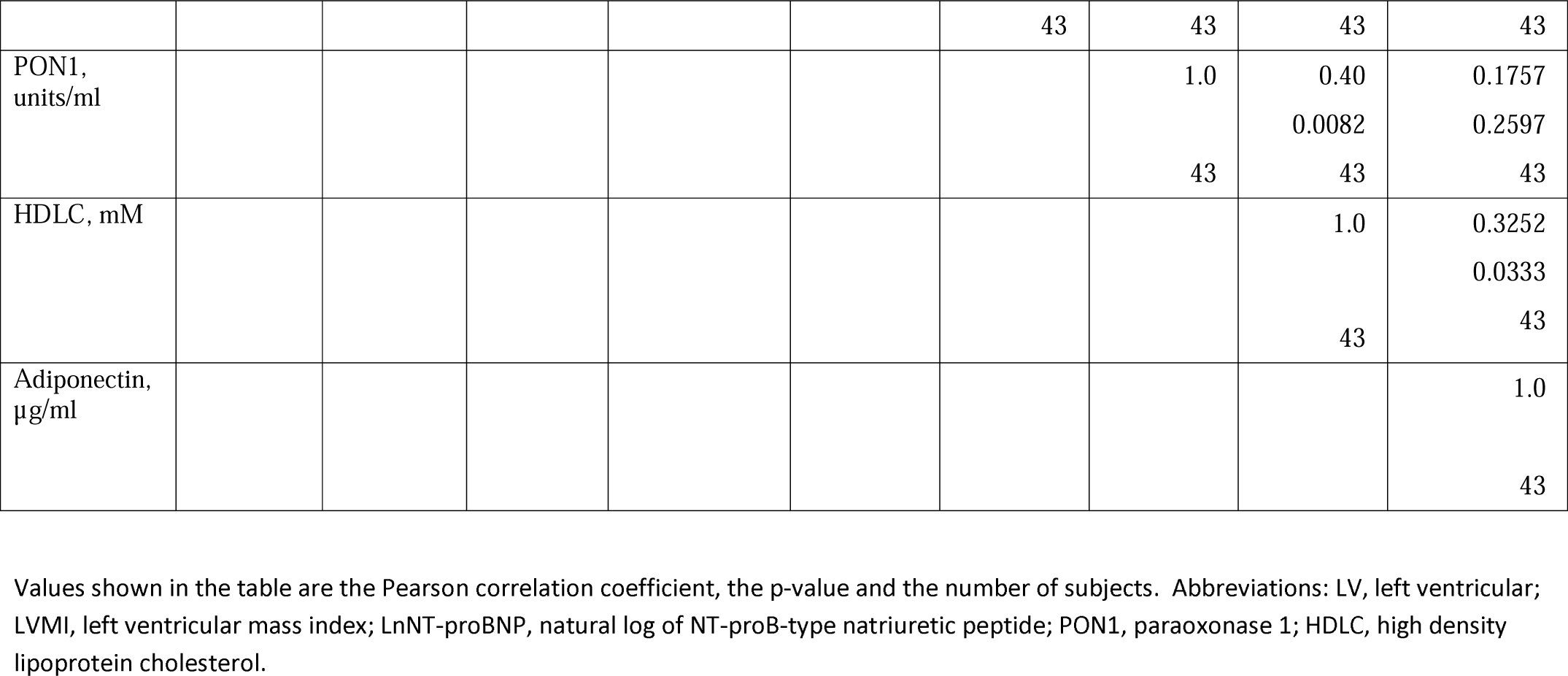
Pearson correlation of change in left ventricular measurements and hemoglobin with PON1, HDL cholesterol, and adiponectin for subjects on dialysis.

For the kidney transplant group, the change in PON1 mass and arylesterase were highly significantly correlated (p<0.0001). Of importance for this study, change in hemoglobin was positively correlated with change on PON1 mass (p=0.0042), but not PON1 activity (p=0.2751). Further, the change in PON1 mass was inversely correlated with the change in left ventricular diastolic volume (p=0.0146), systolic volume (p=0.0114), and LnNT-proBNP (p=0.0062). Although the change in PON1 mass was correlated with the change in HDL cholesterol concentration (p=0.0562), the change in HDL cholesterol was not significantly correlated with left ventricular volume or mass. The change in HDL cholesterol was significantly correlated with the change in PON1 arylesterase (p=0.0003) and adiponectin (p=0.0062).

The median percent change in PON1 mass was −3.9% versus 20.3% for the dialysis and transplant groups, respectively. The change in PON1 mass was not correlated with the average PON1 mass (p=0.56), indicating that baseline PON1 concentrations were not a determinant of the response to kidney transplantation. Thus, the incremental increase in PON1 mass was similar across the PON1 concentration range.

For the dialysis group, the changes primarily reflect biologic variation, as only the change in LnNT-proBNP was significantly different from the null (p=0.0487). The correlation between PON1 mass and arylesterase activity was significant (p <0.0001). Consistent with these two measures having different determinants, PON1 arylesterase variation correlated inversely with the change in left ventricular diastolic volume (p=0.0195) and LnNT-proBNP (p=0.0453), whereas there was no correlation for the change in PON1 mass (p=0.9981, 0.3421, respectively).

PON1 Q192R phenotype frequency was not different between dialysis and transplant subjects (Supplemental Table 1). Further, the Q192R phenotype was not a significant factor for the change in PON1 mass (data not shown).

## Discussion

We have shown for the first time that the increase in PON1 mass post-kidney transplant negatively correlates with the change in left ventricular volume and positively correlates with the change in hemoglobin. Further, we have documented that the change in PON1 mass post-kidney transplant is independent of C-reactive protein, a marker of inflammation. The change in PON1 mass is also independent of the baseline concentration of PON1. The separation of the change in PON1 from changes in CRP, HDL cholesterol, and albumin would be consistent with specific effects that link PON1, hemoglobin, and left ventricular volume in CKD states. PON1 arylesterase did not uniquely share a common factor with the change in hemoglobin or left ventricular end-volume.

Several candidates are suggested by the current literature to explain these relationships. PON1 is involved, as a lactonase, in the metabolic conversion of 5,6-epoxyeicosatrienoic acid (5,6-EET), an epoxy fatty acid that is not a substrate for soluble epoxide hydrolase, and which spontaneously forms a lactone [19-21]. It has been reported that 5,6-EET regulates blood pressure [21]. The epoxyeicosatrienoic acids are also among the molecules that regulate response to mechanotransduction as regulators of TRPV4 [22, 23]. Independently, the production of B-type natriuretic peptide, as reflected by the inverse relationship of PON1 with NT-proBNP, could indicate a link between PON1 and mechano-sensing pathways. Thus, one could speculate that PON1 mass reflects a role in response to left ventricular volume. The cause and effect relationship among changes in left ventricular volume hemoglobin and PON1 mass cannot, however, be elucidated from the current study design.

PON1 has a well-recognized labile metal-binding site, in addition to its enzymatic activity. This labile site requires calcium for enzyme activity, but it can also bind metals. Thus, one potential function of PON1 is in the regulation of iron, zinc, and other divalent cations [24-26]. Rahimi-Ardabili et al. [27] reported that 61 days of 100 mg zinc sulfate increased HDL cholesterol, apoAI, and paraoxonase activity in patients on hemodialysis. Thus, while there is a clear response to zinc, it was not specific to paraoxonase and may be a general effect on HDL production or clearance. This putative function would be dependent upon the mass of PON1 and would have the potential not only to be independent of the lactonase activity but also to be inverse to such activity.

The positive correlation of the change in PON1 mass with the change in hemoglobin could indicate that factors involved in iron metabolism regulate PON1 gene expression in concert with regulating hemoglobin concentration. This would be consistent with an increase in PON1 in response to erythropoietin therapy that has been reported in a single study [28, 29]. However, currently, there is no known link between the regulation of human PON1 gene expression and iron metabolism.

Kidney transplant recipients benefit markedly from their improved kidney function compared to remaining on dialysis, experiencing improved long-term cardiovascular morbidity and mortality (29). While improvement in left ventricular volume and hemoglobin are well-known effects of kidney transplantation, the nature of their relationship to these improved cardiovascular outcomes remains unestablished. In this study, we have shown that PON1 mass and activity may be key mediators in the pathway linking restored kidney function and improved cardiovascular parameters and outcomes. In the future, interventions selectively directed toward increasing PON1 mass may help further understand the mechanisms that connect the kidney and heart in patients with pathological cardiorenal syndromes.

## Data Availability

The data is available upon request.

## Author’s Contributions

PWC, ATY, MMN participated in the research design, funding, performance of the research, writing, and data analysis. RW, CL, LG participated in the research design, performance of the reseach and manuscript revision. GRK participated in performance of the research and data analysis. GVRP coordinated the research design, funding, performance of the research, writing and data analysis.

## Abbreviations

CKD: chronic kidney disease
CMR: cardiac magnetic resonance imaging
ESKD: end-stage kidney disease
HD: hemodialysis
HDL: high density lipoprotein
LV: left ventricular
NT-proBNP: NT pro-B type natriuretic peptide
PON1: paraoxonase 1
PD: peritoneal dialysis

